# Uromodulin as a protein and genetic biomarker for hypertension: a detailed systematic review & meta-analysis

**DOI:** 10.64898/2025.12.29.25343116

**Authors:** Hash Brown Taha, Nandhana Vivek, Samuel Ogunsanya, Haroutyun Joulfayan, Aleksander Bogoniewski, Brian Havens

## Abstract

**Background:** Hypertension (HTN) is a major global contributor to cardiovascular morbidity and mortality, and improved biomarkers are needed to better characterize individual risk, disease progression, and treatment response. Uromodulin (UMOD), the most abundant protein in human urine reflects tubular health and renal sodium handling. Genome-wide association studies (GWAS) have identified common UMOD promoter polymorphisms associated with HTN across diverse populations, sparking interest in UMOD as both a protein and genetic biomarker. We conducted a systematic review and meta-analysis to evaluate the diagnostic, prognostic, and predictive value of UMOD in HTN.

**Methods:** We conducted a systematic review and meta-analysis using PubMed and Embase. We included human and animal studies that evaluated UMOD protein level or UMOD-related GWAS outcomes in relation to HTN or HTN-related outcomes. Risk of bias was assessed using the Newcastle-Ottawa Scale. Inverse variance weighed random-effects models were used to analyze continuous outcomes. Publication bias was evaluated using Begg and Egger tests and funnel plots.

**Results:** Forty studies met the inclusion criteria. Across diagnostic, prognostic, and predictive biomarker studies, 10,726 individuals with HTN and 8,762 normotensive (NTN) participants were included. Furthermore, there were 3,461 women with HTN and 3,605 NTN. GWAS studies included >450,000 HTN and >150,000 NTN individuals. Despite supportive findings from *in vivo* models, across diagnostic, prognostic, predictive, and GWAS qualitative and quantitative analyses, UMOD did not demonstrate clinically meaningful utility as a biomarker for HTN or HTN-related outcomes.

**Conclusion:** Despite strong biological plausibility linking UMOD to renal sodium handling and blood pressure regulation, our meta-analysis demonstrates that UMOD is not a clinically useful protein or genetic biomarker for HTN or HTN-related outcomes.

## Introduction

Hypertension (HTN), defined as a chronic elevation of arterial blood pressure, is a major modifiable cause of cardiovascular disease (CVD) worldwide. Through sustained pressure overload, HTN induces endothelial dysfunction, arterial stiffening, vascular remodeling, and left ventricular hypertrophy, thereby driving the development of major CVDs including coronary artery disease, heart failure, stroke, and peripheral artery disease^1^. CVD remains the leading global cause of morbidity and mortality, affecting more than 523 million individuals worldwide and accounting for nearly 18 million deaths annually^2,3^.

Despite its central role in CVD pathogenesis, HTN is a heterogeneous condition with substantial inter-individual variability in onset, progression, and response to therapy. Conventional blood pressure measurements capture disease severity at a single time point but provide limited insight into the underlying vascular, renal, and neurohormonal mechanisms that determine whether HTN will remain stable, worsen, or respond to intervention. Consequently, there is growing interest in functional biomarkers that reflect endothelial function, arterial compliance, neurohormonal activation, inflammation, and end-organ stress, as these measures may enable earlier risk stratification, distinguish individuals with high-versus low-risk hypertensive phenotypes, and improve prediction of disease progression and cardiovascular outcomes.

Uromodulin (UMOD), also known as Tamm-Horsfall protein, is the most abundant protein in mammalian urine^4^. It is produced exclusively by epithelial cells of the thick ascending limb of the loop of Henle and is primarily secreted into the tubular lumen^5-7^. A smaller fraction enters the bloodstream via basolateral secretion and circulates at lower concentrations^7-10^. UMOD has been extensively used as a noninvasive biomarker in kidney research and clinical nephrology. UMOD urinary levels reflect tubular integrity and are markedly reduced in chronic kidney disease (CKD). In circulation, it also appears to modulate immune responses both locally and systemically^10,11^. UMOD levels have been shown to correlate inversely with inflammatory states and systemic illness^12,13^ as well as diabetes^14,15^ and metabolic syndrome^16^.

Importantly, over a dozen genome-wide association studies and genetic analyses have linked UMOD single-nucleotide polymorphisms (SNPs) to HTN in both humans and mouse models across diverse populations^17-20^. As such, we have seen a growing interest in UMOD as both a protein and genetic biomarker in HTN or HTN-related outcomes. Yet there is currently no comprehensive synthesis integrating protein-level, genetic, and experimental evidence to evaluate the clinical utility of UMOD across diagnostic, prognostic, and predictive contexts in ITN To address this, we performed a systematic review of available studies investigating UMOD in the context of HTN in humans and mice, and quantitatively synthesized relevant data using random-effects meta-analyses to derive robust, evidence-based conclusions regarding the role of UMOD in hypertensive disease

## Methods

We conducted a systematic review, meta-analysis, and meta-regression in accordance with the Preferred Reporting Items for Systematic Reviews and Meta-Analyses (PRISMA) guidelines. This study exclusively analyzed anonymized, previously published data and did not involve direct research on human participants or the collection of personal information; therefore, ethical approval was not required. The study protocol was not registered.

### Data sources and search strategy

We performed a comprehensive and intentionally broad literature search to identify studies investigating UMOD in the context of HTN. Searches were conducted in PubMed and Embase from database inception through December 13th, 2025, using predefined terms related to UMOD, HTN, blood pressure (BP) regulation, and genetic variation (see **Table S1** for full terms). Three independent reviewers (NV, HJ & HBT) screened titles, abstracts, and full-text manuscripts for eligibility. Reference lists of included studies were manually reviewed to identify additional relevant articles, and supplementary searches were conducted using Google Scholar to capture studies not indexed in the primary databases. Any discrepancies in study selection were resolved through consensus discussion.

### Eligibility criteria

We employed a comprehensive and intentionally broad search strategy centered on UMOD and HTN, designed to capture the full scope of relevant evidence. We included all study designs across human and mouse models, encompassing genetic association studies (e.g., UMOD single nucleotide polymorphisms evaluated in hypertensive populations, normotensive controls, or longitudinal cohorts assessing incident HTN risk), protein-based investigations measuring circulating or urinary UMOD in relation to blood pressure or hypertensive status, and mechanistic or experimental studies linking UMOD to blood pressure regulation or hypertensive phenotypes. Studies were eligible for inclusion as long as they examined UMOD in conjunction with HTN or BP-related outcomes, whether HTN was assessed as a primary phenotype, a stratifying variable, a predictor of future disease, or a downstream outcome. All studies meeting inclusion criteria were retained for qualitative synthesis, and those reporting extractable quantitative data were included in random-effects meta-analyses.

### Data extraction

Data from all eligible studies were extracted independently by at least two investigators, with cross-verification performed to ensure accuracy and completeness. When required, summary statistics were transformed using established methods to enable quantitative synthesis. From one study^21^, we could only access the abstract, which included all information relevant for inclusion in the meta-analysis.

The standard error of the mean (SEM) was converted to standard deviation (SD) where required. For studies reporting UMOD levels as medians with interquartile ranges (IQRs), means were estimated as the average of the first quartile, median, and third quartile, and SDs were estimated by dividing the IQR by 1.35. For studies with small sample sizes (n < 25) reporting medians with ranges, means were estimated as the sum of the minimum value, twice the median, and the maximum value divided by four, and SDs were estimated as one quarter of the range. For studies reporting 95% confidence intervals without SDs, SDs were derived from the confidence interval width and sample size. In one proteomics study, UMOD levels were derived from technical replicate measurements and used for quantitative analysis. In studies reporting log-transformed UMOD values, data were back-transformed to the original scale prior to pooling.

### Risk of bias assessment

Risk of bias was assessed using the Newcastle–Ottawa Scale (NOS) for cross-sectional, cohort or case-control studies^22^. Studies were categorized according to the context in which UMOD was evaluated, rather than the original purpose of cohort recruitment. Specifically, classification was based on whether UMOD measurement occurred cross-sectionally, longitudinally, or was used in a prognostic or predictive framework. As such, assignment to cross-sectional, cohort or case-control categories reflects the timing and intended use of the UMOD biomarker (e.g., diagnostic versus prognostic), and not necessarily the primary design or objectives of the parent study.

### Statistical analysis

Random-effects meta-analyses were performed using R software (version 2024.12.0+467) with the *metafor* package. An inverse-variance weighting approach was applied to account for both within-study and between-study variability. For continuous outcomes, pooled standardized mean differences (SMDs) with 95% confidence intervals (CIs) were calculated using Cohen’s *d* to enable comparison across studies reporting UMOD levels on different scales. Cohen’s *d* and pooled standard deviation were calculated using standard previously published formula^23^.

Publication bias^24^ was assessed using Begg’s rank correlation test, Egger’s regression test, and Deeks’ funnel plot asymmetry test, complemented by visual inspection of funnel plots. All publication bias analyses were conducted using R software (version 2024.12.0+467).

## Results

Following full-text screening, a total of 40 studies^17-21,25-59^ met the inclusion criteria and were included in the systematic review (**Figure 1**). Of these, 15 studies (**Table 1**) evaluated UMOD in a cross-sectional and diagnostic context, with 13 contributing quantitative data to the meta-analysis. Nine studies assessed the prognostic value of UMOD (**Table 2**), and three studies examined UMOD as a predictive biomarker for HTN-related outcomes (**Supplementary Table 2**). In addition, 16 genome-wide association studies (GWAS) investigated UMOD genetic variation (**Table 3**), of which ten were included in meta-analyses, while two Mendelian randomization (MR) studies (**Supplementary Table 3**) and five animal studies (**Supplementary Table 4**) were included for qualitative synthesis.

**Figure 1.**
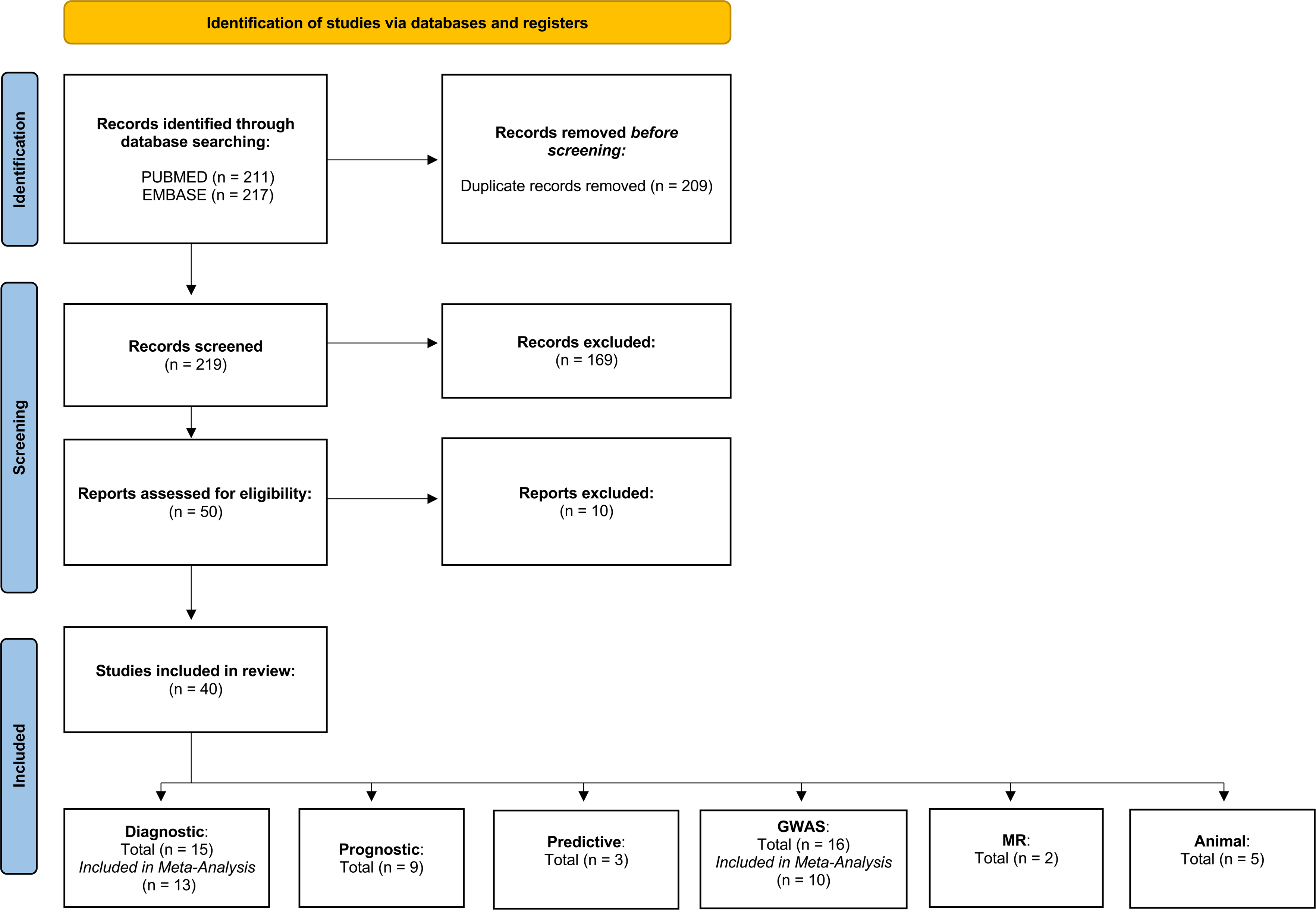
PRISMA flow diagram of study selection. Flow diagram illustrating the identification, screening, eligibility assessment, and inclusion of studies evaluating uromodulin (UMOD) in the context of hypertension. Following database searches and full-text review, 40 studies were included in the systematic review and categorized according to biomarker application, including diagnostic, prognostic, predictive, genome-wide association studies, Mendelian randomization (MR) analyses, and animal studies. Numbers of studies included in qualitative synthesis and quantitative meta-analyses are indicated where applicable.

Across the diagnostic, prognostic, and predictive studies, a total of 10,726 individuals with HTN and 8,762 NTN were evaluated. The majority of these studies focused on essential hypertension, while six studies examined CKD-HTN. One longitudinal cohort enrolled participant initially free of HTN, CKD, and CVD to assess racial differences in UMOD biology. Among clinical studies, UMOD was primarily quantified in urine (19 studies), with serum or plasma used in 10 studies, and UMOD-positive extracellular vesicles (EVs) assessed in one study. Measurement techniques included enzyme-linked immunosorbent assay (ELISA) in 14 studies, electrochemiluminescence immunoassay (ECLIA) in 4, radioimmunoassay (RIA) in 2, and Western blotting (WB) in 1, while multiplex bead assays (3 studies), flow cytometry (1 study), and LC-MS/MS proteomics (1 study) were applied in selected cohorts. Genetic studies contributed >400,00 HTN and >150,000 NTN participants. These analyses included 6 GWAS exclusively enrolling individuals with HTN, 9 GWAS conducted in mixed HTN/NTN populations, and one GWAS focused on CKD-HTN, using SNP genotyping, targeted allele assays, and large-scale biobank data to investigate UMOD genetic variation across diverse populations. Detailed information for all studies can be found in **Tables 1-4** and **Supplementary Tables 2-4**.

### Risk of bias

Studies were classified as cross-sectional, cohort, or case-control designs, and methodological quality was assessed using the Newcastle–Ottawa Scale (NOS). Overall, the majority of the studies exhibited low bias (**Supplementary Figure S1-3**).

### UMOD as a biomarker

#### Diagnostic

Overall, out of the 15 studies assessing UMOD as a cross-sectional biomarker for HTN, only 13 studies had quantitative data. We meta-analyzed UMOD levels in individuals with HTN compared with NTN and pre-HTN controls across four analyses (see **Table 1** for study-specific cohort details): (1) all HTN etiologies versus NTN/Pre-HTN (**Figure 2A**), (2) isolated HTN after exclusion of all comorbid conditions (**Figure 2B**), (3) urine-based UMOD measurements (**Figure 2C**), and (4) blood-based UMOD measurements (**Figure 2D**). Across all analyses, the analyses demonstrated no statistical significance in UMOD levels in HTN vs. NTN. A single study emerged as a major outlier with an extreme effect size and was the primary driver of heterogeneity; aside from this study, effect estimates were small and directionally inconsistent.

**Figure 2.**
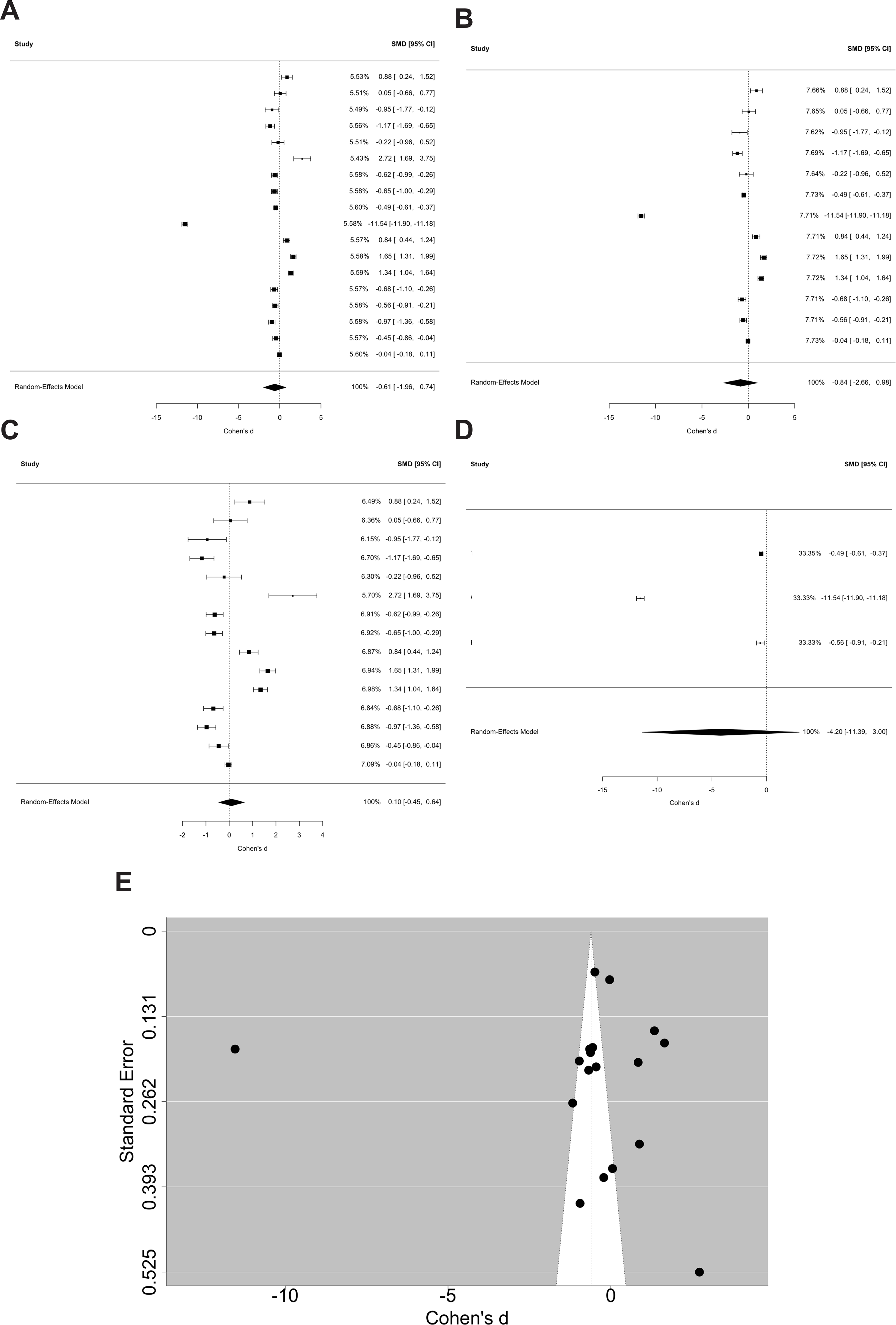
Meta-analysis of uromodulin (UMOD) levels in hypertension (HTN). (A) Forest plot comparing UMOD levels in individuals with hypertension (HTN) of all etiologies, including comorbid conditions, versus normotensive (NTN) or pre-hypertensive (PreHTN) controls. (B) Forest plot restricted to isolated hypertension versus normotension. (C) Subgroup analysis of urinary UMOD measurements. (D) Subgroup analysis of blood-based UMOD measurements (serum or plasma). Effect sizes are shown as standardized mean differences (Cohen’s d) with 95% confidence intervals using random-effects models. Across all analyses, pooled estimates were centered near the null and did not demonstrate a statistically significant difference in UMOD levels between hypertensive and non-hypertensive groups. (E) Funnel plot assessing small-study effects and publication bias; visual symmetry and formal testing (Egger’s regression and Begg’s rank correlation) indicated no evidence of publication bias.

Blood-based analyses showed a weak downward trend in UMOD levels among individuals with, whereas urine-based analyses demonstrated a modest trend in the opposite direction; however, neither pattern was robust or statistically meaningful. Overall, these findings do not support a reproducible difference in UMOD levels in HTN relative to NTN or preHTN. We did not conduct sensitivity analyses due to most studies being insignificant. Visual inspection of the funnel plot demonstrated overall symmetry around the pooled effect estimate, with no clear evidence of small-study effects or directional asymmetry (**Figure 2E**). This was supported by formal statistical testing: Egger’s regression test did not indicate funnel plot asymmetry (t = - 0.27, p = 0.79), and Begg’s rank correlation test likewise showed no evidence of publication bias (Kendall’s τ = -0.15, p = 0.41).

#### Prognostic

We characterized nine studies as prognostic, examining whether circulating or urinary UMOD levels predict incident HTN or BP-related outcomes, including SBP, DBP, and related metrics (see **Table 2**). Overall, prognostic evidence was mixed and matrix dependent. Several studies reported inverse associations between serum UMOD and incident HTN or higher SBP/DBP in univariate models, with effect sizes generally attenuating and frequently losing significance after multivariable adjustment for demographics, kidney function, cardiovascular risk factors, and medication use. Urinary UMOD showed less consistent prognostic value: while some analyses demonstrated inverse associations with contemporaneous BP measures (e.g., DBP, MAP, pulse pressure) or incident HTN in unadjusted or partially adjusted models, most fully adjusted models and longitudinal analyses did not support urinary UMOD as an independent predictor of incident HTN or long-term BP change. Correlation analyses likewise showed weak or null associations. Notably, several studies suggested that lower UMOD levels were more strongly associated with downstream cardiovascular and kidney outcomes (e.g., acute kidney injury, eGFR decline, end-stage kidney disease, and composite cardiovascular events) in people with HTN than with HTN itself. Collectively, these findings indicate that while UMOD may reflect broader cardio-renal risk, its utility as a robust, independent prognostic biomarker for incident HTN or BP trajectories remains limited and inconsistent across biological matrices and levels of adjustment.

#### Predictive

Three studies assessed UMOD responses to dietary salt (high or low). Across these studies (**Supplementary Table 2**), both plasma and urinary UMOD levels decreased with high-salt intake and increased under low-salt conditions, with modest correlations observed between plasma UMOD and contemporaneous SBP and DBP. However, baseline urinary UMOD levels did not predict subsequent BP changes following salt interventions, and associations with blood pressure change were weak and non-significant after multivariable adjustment, indicating that UMOD primarily reflects acute salt-related physiological responses rather than serving as a robust predictor of salt sensitivity-induced BP changes.

#### UMOD SNPs

Overall, out of the 16 studies assessing UMOD SNPs in HTN, only ten (**Table 3**) studies had quantitative data included in the meta-analysis. Homozygous major allele carriers were compared with individuals carrying at least one minor allele (heterozygous or homozygous minor). For urinary UMOD levels (**Figure 3A**), the pooled random effects estimate indicated a small difference between genotype groups (SMD = 0.19, 95% CI 0.00–0.39, p = 0.047). However, SNP stratified effect estimates were modest and largely overlapped with the null, with no single variant demonstrating a consistent association across studies. Analysis of SBP (**Figure 3B**) and DBP (**Figure 3C**) demonstrated wide confidence intervals and no statistically significant pooled difference between genotype groups.

**Figure 3.**
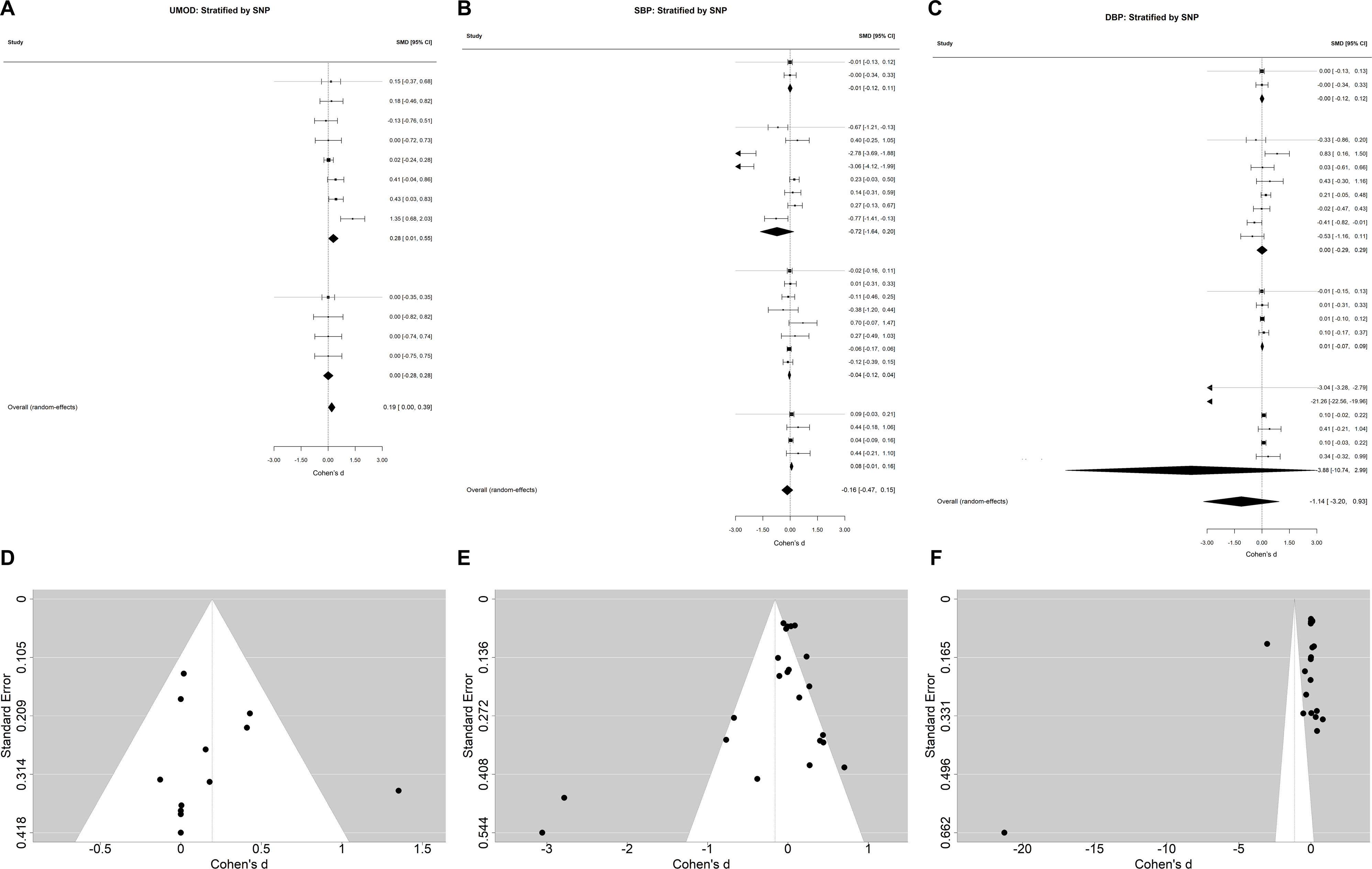
Meta-analysis of UMOD associated genetic variants and quantitative phenotypes for individuals carrying at least one minor allele (heterozygous or homozygous minor), stratified by individual UMOD-associated single-nucleotide polymorphisms (SNPs). **(A)** Forest plot of standardized mean differences (SMDs) for UMOD levels stratified by UMOD promoter SNP genotype. **(B)** Forest plot of systolic blood pressure (SBP) stratified by UMOD promoter SNP genotype. **(C)** Forest plot of diastolic blood pressure (DBP) stratified by UMOD promoter SNP genotype. Effect sizes are shown as SMDs with 95% confidence intervals using random-effects models. Across all SNP-stratified analyses, pooled effect estimates were centered near the null and did not demonstrate statistically significant associations between UMOD promoter variants and UMOD levels, SBP, or DBP. (**D–F**) Funnel plots corresponding to panels **A–C**, respectively, assessing small-study effects and publication bias; visual inspection demonstrated overall symmetry, with little evidence of publication bias using Begg’s rank correlation or Egger’s regression.

Visual inspection of the funnel plot for urinary UMOD suggested overall symmetry around the pooled effect estimate, with no clear evidence of small-study effects (**Figure 3D**), supported by Egger’s regression (z = 0.29, p = 0.77) and Begg’s rank correlation (τ = 0.15, p = 0.55). In contrast, SBP analyses showed evidence of funnel plot asymmetry (**Figure 3E**) by Egger’s regression (z = -2.57, p = 0.010), though Begg’s test did not indicate significant asymmetry (τ = −0.10, p = 0.54) (**Figure 3D**). For DBP, both Egger’s regression (z = -3.63, p < 0.001) and Begg’s rank correlation (τ = -0.36, p = 0.028) suggested funnel plot asymmetry (**Figure 3F**). Given the limited number of contributing studies, small genotype subgroup sizes, and substantial between-study heterogeneity, these patterns are more consistent with statistical imprecision than with a reproducible genetic effect. We did not conduct sensitivity analyses due to the small number of included studies.

#### MR

Two independent MR studies provided supporting evidence that higher UMOD levels, driven by UMOD gene variants, exerted potential causal effects on BP regulation (**Supplementary Table S4**). In both studies, genetic instruments for increased UMOD were consistently associated with higher SBP, higher DBP, and increased risk of HTN.

In the first study, urinary UMOD was evaluated using the IVW approach, while serum UMOD was assessed using IVW, MR-Egger, weighted median, and MR-PRESSO. These analyses identified a causal association between genetically elevated serum UMOD and increased DBP, with consistent effect estimates across datasets. In parallel, IVW analyses demonstrated that higher genetically predicted urinary UMOD increased SBP, DBP, and incident HTN, supporting urinary UMOD as a causal risk factor for elevated BP and incident HTN. The second study examined the causal relationships among UMOD, CKD and BP. One-sample MR using the UMOD polymorphism rs12917707 demonstrated that genetically higher urinary UMOD was strongly associated with a decline in eGFR, indicating a detrimental effect on kidney function. These findings were reinforced by a two-sample Mendelian randomization analysis across four large GWAS datasets, encompassing CKD risk and BP traits. Genetically increased urinary UMOD was associated with lower eGFR, increased odds of CKD, and higher SBP and DBP, supporting a causal role for UMOD in both renal dysfunction and HTN.

#### Race/Ethnicity

Four studies evaluated differences in UMOD levels by race and their relationship to HTN and together demonstrated that UMOD is lower in Black participants compared with White participants, with ancestry-specific genetic associations. In the CARDIA cohort, middle-aged Black participants had lower urinary UMOD concentrations than White participants after adjustment for age, sex, and urine creatinine, with similar effect estimates after full multivariable adjustment. A multi-ethnic urine proteomics study comparing Sub-Saharan African and European cohorts supported these findings, showing that 3 of 4 measured UMOD peptide fragments were reproducibly lower in Black participants across multiple replications. In contrast to biomarker differences, the genetic contribution of UMOD to HTN differed by ancestry and was not evaluated due to potential confounding by regional differences in the distribution of risk factors, such as access to healthcare, and age. In the Million Veteran Program, the UMOD promoter variant rs4293393 was associated with increased UMOD expression and was significantly associated with HTN in non-Hispanic White participants but showed no association in non-Hispanic Black participants. In the African American Study of Kidney Disease and Hypertension, which enrolled Black adults with HTN-related kidney disease, lower serum UMOD levels reflected worse underlying tubular health, with a 50% lower baseline UMOD level independently associated with a higher risk of kidney failure, and each 1-SD steeper annual decline in UMOD associated with a 67% increased risk of kidney failure. These findings combined may suggest that Black populations exhibit lower overall UMOD levels and experience weaker adverse effects in relation to commonly studied UMOD HTN risk variants.

#### Animal

Experimental investigations from five studies^20,32,52,58,59^ have utilized altered U*mod* mice models, hypertensive pregnant rat strains, and felines to determine the role of Umod in renal physiology and BP regulation (see Supplementary **Table S4** for full details).

In a seminal study, Graham et al.^20^ demonstrated that UMOD plays a key role in regulating blood pressure and salt sensitivity by comparing wild type (*Umod*+/+) and Umod knockout (*Umod*-/-) mice. They showed that *Umod* deficient mice had significantly lower SBP at baseline and were protected from salt induced HTN during high salt intake. This BP protection in *Umod* knockout mice was accompanied by increased urinary sodium excretion, higher urine volume, and a leftward shift in renal pressure natriuresis curves, indicating enhanced sodium handling. Mechanistically, loss of *Umod* increased TNF-alpha activity in the thick ascending limb, leading to reduced NKCC2 expression, decreased sodium reabsorption, lower extracellular volume, and ultimately reduced BP. Similar findings were found in another murine study^32^.

Follow-up studies in rat models have investigated hypertensive pregnancy, using SHRSP and WKY strains. The first study^59^ emphasized changes in renal UMOD protein and peptide levels, including shifts in polymerization states that were sensitive to nifedipine treatment as indicated by a shift in phosphorylation. From the same group, the most recent study^52^ linked pregnancy-associated UMOD alterations to renal injury markers NGAL and KIM-1 independent of SBP demonstrating convergence on renal damage between mice and rat models with species- and context-dependent effects. An additional study in geriatric cats^58^ examined *Umod* SNPs across multiple breeds, revealing associations with SBP.

## Discussion

In this study, we included 40 studies evaluating UMOD in the context of HTN, comprising 15 cross-sectional/diagnostic studies (13 contributing quantitative data), 9 prognostic studies, 3 predictive studies, 16 GWAS (10 included in meta-analyses), 2 MR studies, and 5 animal studies. Across diagnostic, prognostic, and predictive analyses, more than 10,700 individuals with HTN and 8,700 NTN were evaluated, while genetic analyses included over 650,000 participants. Studies spanned diverse populations, biological matrices (urine, serum, plasma), and analytical platforms, and overall risk of bias was low based on NOS assessment.

Across all human analyses, UMOD did not demonstrate clinically meaningful utility as a biomarker for HTN. Meta-analyses showed no reproducible differences in UMOD levels between HTN and NTN or preHTN, with isolated significant findings driven by single outlier studies rather than consistent effects. Prognostic associations between baseline UMOD and incident HTN or longitudinal BP measures were inconsistent and frequently attenuated after multivariable adjustment, while predictive analyses showed that UMOD responds acutely to dietary salt manipulation but does not reliably predict salt sensitivity-related BP changes. GWAS meta-analyses demonstrated effect estimates centered near the null for UMOD levels, SBP, and DBP, despite evidence of funnel plot asymmetry in some BP analyses likely driven by small study numbers and heterogeneity. In contrast, MR studies and animal models consistently supported a causal role for UMOD in renal sodium handling and BP regulation. Collectively, these findings highlight a clear disconnect between mechanistic and genetic evidence and the limited clinical utility of UMOD as a biomarker for HTN.

Additionally, we are aware of one large prospective study that measured serum UMOD levels in a community-based cohort and reported that higher UMOD quantiles were associated with lower risks of all-cause mortality and incident CVD^60^. Notably, these associations were observed in an older general population and were not evaluated specifically in individuals with HTN. Thus, while our findings indicate that UMOD is not a useful diagnostic, prognostic, or predictive biomarker for HTN, UMOD may still be relevant to cardiovascular and renal outcomes more broadly, including CKD^61^. However, given the very limited number of studies examining UMOD in relation to CVD, additional well-powered prospective studies are needed to clarify its role outside the context of HTN.

Only one included study assessed UMOD in EVs, limiting conclusions regarding EV-associated UMOD in HTN. EVs represent a heterogeneous population of membrane-bound particles, including exosomes, ectosomes), and apoptotic bodies, and have been extensively studied as biomarkers and mediators across a wide range of metabolic^62^, cardiovascular^63^, neurological^64-66^, and oncologic^67^ disorders. Because EV cargo reflects cell-type- and state-specific signaling, EV-associated UMOD may capture biologically meaningful information that is not detectable in bulk urine or circulation and could help contextualize otherwise modest or inconsistent changes in total UMOD levels. Future studies specifically quantifying UMOD within well-characterized EV subpopulations may therefore provide insight into whether EV-associated UMOD better reflect biomarker changes are physiologically relevant.

In conclusion, despite strong biological plausibility and supportive evidence from animal and genetic studies, comprehensive synthesis of human data indicates that UMOD is not a clinically useful diagnostic, prognostic, or predictive biomarker for HTN.

## Data Availability

All data produced in the present study are available upon reasonable request to the authors

## Authorship contributions

BH proposed investigating UMOD in the context of cardiovascular disease, while HBT subsequently refined the focus to hypertension and conceptualized and designed all aspects of the study. NV, HBT, HJ & BH performed abstract and full-text screening. NV, HBT & SO collected the data. HBT & NV analyzed the data. HBT, NV, SO & AB wrote the manuscript. HBT supervised and managed the research team.

## References

1 Fuchs, F. D. & Whelton, P. K. High Blood Pressure and Cardiovascular Disease. Hypertension 75, 285–292 (2020). 10.1161/HYPERTENSIONAHA.119.14240

2 Nedkoff, L., Briffa, T., Zemedikun, D., Herrington, S. & Wright, F. L. Global Trends in Atherosclerotic Cardiovascular Disease. Clin Ther 45, 1087–1091 (2023). 10.1016/j.clinthera.2023.09.020

3 Roth, G. A. et al. Global Burden of Cardiovascular Diseases and Risk Factors, 1990-2019: Update From the GBD 2019 Study. J Am Coll Cardiol 76, 2982–3021 (2020). 10.1016/j.jacc.2020.11.010

4 Lhotta, K. Uromodulin and chronic kidney disease. Kidney Blood Press Res 33, 393-398 (2010). 10.1159/000320681

5 Cavallone, D., Malagolini, N. & Serafini-Cessi, F. Mechanism of release of urinary Tamm-Horsfall glycoprotein from the kidney GPI-anchored counterpart. Biochem Biophys Res Commun 280, 110–114 (2001). 10.1006/bbrc.2000.4090

6 Schaeffer, C., Santambrogio, S., Perucca, S., Casari, G. & Rampoldi, L. Analysis of uromodulin polymerization provides new insights into the mechanisms regulating ZP domain-mediated protein assembly. Mol Biol Cell 20, 589–599 (2009). 10.1091/mbc.e08-08-0876

7 Devuyst, O., Olinger, E. & Rampoldi, L. Uromodulin: from physiology to rare and complex kidney disorders. Nat Rev Nephrol 13, 525–544 (2017). 10.1038/nrneph.2017.101

8 Scherberich, J. E. et al. Serum uromodulin-a marker of kidney function and renal parenchymal integrity. Nephrol Dial Transplant 33, 284–295 (2018). 10.1093/ndt/gfw422

9 Steubl, D. et al. Plasma Uromodulin Correlates With Kidney Function and Identifies Early Stages in Chronic Kidney Disease Patients. Medicine (Baltimore) 95, e3011 (2016). 10.1097/MD.0000000000003011

10 El-Achkar, T. M. et al. Tamm-Horsfall protein translocates to the basolateral domain of thick ascending limbs, interstitium, and circulation during recovery from acute kidney injury. Am J Physiol Renal Physiol 304, F1066–1075 (2013). 10.1152/ajprenal.00543.2012

11 El-Achkar, T. M. et al. Tamm-Horsfall protein protects the kidney from ischemic injury by decreasing inflammation and altering TLR4 expression. Am J Physiol Renal Physiol 295, F534–544 (2008). 10.1152/ajprenal.00083.2008

12 Micanovic, R. et al. Tamm-Horsfall Protein Regulates Granulopoiesis and Systemic Neutrophil Homeostasis. J Am Soc Nephrol 26, 2172–2182 (2015). 10.1681/ASN.2014070664

13 Micanovic, R., LaFavers, K., Garimella, P. S., Wu, X. R. & El-Achkar, T. M. Uromodulin (Tamm-Horsfall protein): guardian of urinary and systemic homeostasis. Nephrol Dial Transplant 35, 33–43 (2020). 10.1093/ndt/gfy394

14 Then, C. et al. Serum Uromodulin Is Associated With But Does Not Predict Type 2 Diabetes in Elderly KORA F4/FF4 Study Participants. J Clin Endocrinol Metab 104, 3795–3802 (2019). 10.1210/jc.2018-02557

15 Leiherer, A. et al. Serum uromodulin is associated with impaired glucose metabolism. Medicine (Baltimore) 96, e5798 (2017). 10.1097/MD.0000000000005798

16 Then, C. et al. Serum uromodulin is inversely associated with the metabolic syndrome in the KORA F4 study. Endocr Connect 8, 1363–1371 (2019). 10.1530/EC-19-0352

17 Soltesz, B. et al. The genetic risk for hypertension is lower among the Hungarian Roma population compared to the general population. PLoS One 15, e0234547 (2020). 10.1371/journal.pone.0234547

18 Villagomez Fuentes, L. E., et al. Effect of a common UMOD variant on kidney function, blood pressure, cognitive and physical function in a community-based cohort of older adults. J Hum Hypertens 36, 983–988 (2022). 10.1038/s41371-021-00608-2

19 Akwo, E. A. et al. Phenome-Wide Association Study of UMOD Gene Variants and Differential Associations With Clinical Outcomes Across Populations in the Million Veteran Program a Multiethnic Biobank. Kidney Int Rep 7, 1802–1818 (2022). 10.1016/j.ekir.2022.05.011

20 Graham, L. A. et al. Validation of uromodulin as a candidate gene for human essential hypertension. Hypertension 63, 551–558 (2014). 10.1161/HYPERTENSIONAHA.113.01423

21 Morshed, M. R. et al. Evaluation of Early Renal Involvement in Essential Hypertension by Measuring Urinary Biomarkers. Mymensingh Med J 31, 1183–1191 (2022).

22 Stang, A. Critical evaluation of the Newcastle-Ottawa scale for the assessment of the quality of nonrandomized studies in meta-analyses. Eur J Epidemiol 25, 603–605 (2010). 10.1007/s10654-010-9491-z

23 Taha, H. B. & Ati, S. S. Evaluation of alpha-synuclein in CNS-originating extracellular vesicles for Parkinsonian disorders: A systematic review and meta-analysis. CNS Neurosci Ther 29, 3741–3755 (2023). 10.1111/cns.14341

24 Lin, L. & Chu, H. Quantifying publication bias in meta-analysis. Biometrics 74, 785–794 (2018). 10.1111/biom.12817

25 Dulawa, J., Kokot, F., Kokot, M. & Pander, H. Urinary excretion of Tamm-Horsfall protein in normotensive and hypertensive elderly patients. J Hum Hypertens 12, 635–637 (1998). 10.1038/sj.jhh.1000680

26 Torffvit, O., Agardh, C. D. & Thulin, T. A study of Tamm-Horsfall protein excretion in hypertensive patients and type 1 diabetic patients. Scand J Urol Nephrol 33, 187–191 (1999). 10.1080/003655999750015970

27 Torffvit, O., Melander, O. & Hulten, U. L. Urinary excretion rate of Tamm-Horsfall protein is related to salt intake in humans. Nephron Physiol 97, p31–36 (2004). 10.1159/000077600

28 Kottgen, A. et al. Uromodulin levels associate with a common UMOD variant and risk for incident CKD. J Am Soc Nephrol 21, 337–344 (2010). 10.1681/ASN.2009070725

29 Iwai, N., Kajimoto, K., Kokubo, Y. & Tomoike, H. Extensive genetic analysis of 10 candidate genes for hypertension in Japanese. Hypertension 48, 901–907 (2006). 10.1161/01.HYP.0000242485.23148.bb

30 Padmanabhan, S. et al. Genome-wide association study of blood pressure extremes identifies variant near UMOD associated with hypertension. PLoS Genet 6, e1001177 (2010). 10.1371/journal.pgen.1001177

31 Han, J. et al. Common variants of the UMOD promoter associated with blood pressure in a community-based Chinese cohort. Hypertens Res 35, 769–774 (2012). 10.1038/hr.2012.51

32 Trudu, M. et al. Common noncoding UMOD gene variants induce salt-sensitive hypertension and kidney damage by increasing uromodulin expression. Nat Med 19, 1655–1660 (2013). 10.1038/nm.3384

33 Matafora, V. et al. Quantitative proteomics reveals novel therapeutic and diagnostic markers in hypertension. BBA Clin 2, 79–87 (2014). 10.1016/j.bbacli.2014.10.001

34 Jian, L., Fa, X., Zhou, Z. & Liu, S. Functional analysis of UMOD gene and its effect on inflammatory cytokines in serum of essential hypertension patients. Int J Clin Exp Pathol 8, 11356–11363 (2015).

35 Troyanov, S. et al. Clinical, Genetic, and Urinary Factors Associated with Uromodulin Excretion. Clin J Am Soc Nephrol 11, 62–69 (2016). 10.2215/CJN.04770415

36 Algharably, E. A. H. et al. Uromodulin associates with cardiorenal function in patients with hypertension and cardiovascular disease. J Hypertens 35, 2053–2058 (2017). 10.1097/HJH.0000000000001432

37 Nqebelele, N. U., Dickens, C., Dix-Peek, T., Duarte, R. & Naicker, S. Urinary Uromodulin Levels and UMOD Variants in Black South Africans with Hypertension-Attributed Chronic Kidney Disease. Int J Nephrol 2019, 8094049 (2019). 10.1155/2019/8094049

38 Santelli, A. et al. Senescent Kidney Cells in Hypertensive Patients Release Urinary Extracellular Vesicles. J Am Heart Assoc 8, e012584 (2019). 10.1161/JAHA.119.012584

39 Steubl, D. et al. Association of serum and urinary uromodulin and their correlates in older adults-The Cardiovascular Health Study. Nephrology (Carlton) 25, 522–526 (2020). 10.1111/nep.13688

40 Then, C. et al. Serum uromodulin is inversely associated with arterial hypertension and the vasoconstrictive prohormone CT-proET-1 in the population-based KORA F4 study. PLoS One 15, e0237364 (2020). 10.1371/journal.pone.0237364

41 Bakhoum, C. Y. et al. The Relationship Between Urine Uromodulin and Blood Pressure Changes: The DASH-Sodium Trial. Am J Hypertens 34, 154–156 (2021). 10.1093/ajh/hpaa140

42 Du, M. F. et al. Associations of plasma uromodulin and genetic variants with blood pressure responses to dietary salt interventions. J Clin Hypertens (Greenwich) 23, 1897–1906 (2021). 10.1111/jch.14347

43 You, R. et al. High Level of Uromodulin Increases the Risk of Hypertension: A Mendelian Randomization Study. Front Cardiovasc Med 8, 736001 (2021). 10.3389/fcvm.2021.736001

44 Ponte, B. et al. Mendelian randomization to assess causality between uromodulin, blood pressure and chronic kidney disease. Kidney Int 100, 1282–1291 (2021). 10.1016/j.kint.2021.08.032

45 Wang, Y. et al. Associations of Serum Uromodulin and Its Genetic Variants With Blood Pressure and Hypertension in Chinese Adults. Front Cardiovasc Med 8, 710023 (2021). 10.3389/fcvm.2021.710023

46 Algharably, E. A. et al. Longitudinal effects of a common UMOD variant on kidney function, blood pressure, cognitive and physical function in older women and men. J Hum Hypertens 37, 709–717 (2023). 10.1038/s41371-022-00781-y

47 Khan, M. B. et al. Associations of Urine Biomarkers of Kidney Tubule Health With Incident Hypertension and Longitudinal Blood Pressure Change in Middle-Aged Adults: The CARDIA Study. Hypertension 80, 1353–1362 (2023). 10.1161/HYPERTENSIONAHA.123.21084

48 Josipovic, J. et al. Uromodulin - a Link between Sodium Excretion and Alteration in Circadian Blood Pressure Pattern in Prehypertensives. Acta Clin Croat 62, 313–322 (2023). 10.20471/acc.2023.62.02.09

49 Chen, T. K. et al. Associations of Baseline and Longitudinal Serum Uromodulin With Kidney Failure and Mortality: Results From the African American Study of Kidney Disease and Hypertension (AASK) Trial. Am J Kidney Dis 83, 71–78 (2024). 10.1053/j.ajkd.2023.05.017

50 Jaques, D. A. et al. Association of serum copeptin and urinary uromodulin with kidney function, blood pressure and albuminuria at 6 weeks post-partum in pre-eclampsia. Front Cardiovasc Med 11, 1310300 (2024). 10.3389/fcvm.2024.1310300

51 Brobak, K. M., et al. Novel biomarkers in patients with uncontrolled hypertension with and without kidney damage. Blood Press 33, 2323980 (2024). 10.1080/08037051.2024.2323980

52 Mary, S. et al. Pregnancy-associated changes in urinary uromodulin excretion in chronic hypertension. J Nephrol 37, 597–610 (2024). 10.1007/s40620-023-01830-6

53 Ikeme, J. C. et al. The Association of Plasma and Urine Uromodulin With Cardiovascular Disease in Persons With Hypertension and CKD. Am J Kidney Dis 84, 799–802 (2024). 10.1053/j.ajkd.2024.05.012

54 McCallum, L. et al. UMOD Genotype-Blinded Trial of Ambulatory Blood Pressure Response to Torasemide. Hypertension 81, 2049–2059 (2024). 10.1161/HYPERTENSIONAHA.124.23122

55 An, D. W. et al. Urinary Proteomics and Systems Biology Link Eight Proteins to the Higher Risk of Hypertension and Related Complications in Blacks Versus Whites. Proteomics 25, e202400207 (2025). 10.1002/pmic.202400207

56 Degenaar, A., Kruger, R., Jacobs, A., Pieters, M. & Mels, C. M. Differential associations between kidney and vascular health biomarkers in young adults stratified by blood pressure status: The African Prospective study on the Early Detection and Identification of Cardiovascular disease and Hypertension study. J Hypertens 43, 1339–1347 (2025). 10.1097/HJH.0000000000004051

57 Ikeme, J. C. et al. Associations of Plasma and Urine Uromodulin With Kidney Disease Progression in Persons With Chronic Kidney Disease and Hypertension: The Systolic Blood Pressure Intervention Trial. Kidney Med 7, 101118 (2025). 10.1016/j.xkme.2025.101118

58 Jepson, R. E., Warren, H. R., Syme, H. M., Elliott, J. & Munroe, P. B. Uromodulin gene variants and their association with renal function and blood pressure in cats: a pilot study. J Small Anim Pract 57, 580–588 (2016). 10.1111/jsap.12582

59 Mary, S. et al. Polymerization-Incompetent Uromodulin in the Pregnant Stroke-Prone Spontaneously Hypertensive Rat. Hypertension 69, 910–918 (2017). 10.1161/HYPERTENSIONAHA.116.08826

60 Steubl, D. et al. Association of serum uromodulin with mortality and cardiovascular disease in the elderly-the Cardiovascular Health Study. Nephrol Dial Transplant 35, 1399–1405 (2020). 10.1093/ndt/gfz008

61 El-Achkar, T. M. & Wu, X. R. Uromodulin in kidney injury: an instigator, bystander, or protector? Am J Kidney Dis 59, 452–461 (2012). 10.1053/j.ajkd.2011.10.054

62 Akbar, N., Azzimato, V., Choudhury, R. P. & Aouadi, M. Extracellular vesicles in metabolic disease. Diabetologia 62, 2179–2187 (2019). 10.1007/s00125-019-05014-5

63 Sahoo, S. et al. Therapeutic and Diagnostic Translation of Extracellular Vesicles in Cardiovascular Diseases: Roadmap to the Clinic. Circulation 143, 1426–1449 (2021). 10.1161/CIRCULATIONAHA.120.049254

64 Taha, H. B. Alzheimer’s disease and related dementias diagnosis: a biomarkers meta-analysis of general and CNS extracellular vesicles. npj Dementia 1 (2025). 10.1038/s44400-024-00002-y

65 Taha, H. B. & Bogoniewski, A. Analysis of biomarkers in speculative CNS-enriched extracellular vesicles for parkinsonian disorders: a comprehensive systematic review and diagnostic meta-analysis. J Neurol 271, 1680–1706 (2024). 10.1007/s00415-023-12093-3

66 Dutta, S., Hornung, S., Taha, H. B. & Bitan, G. Biomarkers for parkinsonian disorders in CNS-originating EVs: promise and challenges. Acta Neuropathol 145, 515–540 (2023). 10.1007/s00401-023-02557-1

67 Semeradtova, A. et al. Extracellular vesicles in cancer s communication: messages we can read and how to answer. Mol Cancer 24, 86 (2025). 10.1186/s12943-025-02282-1

