## Supplementary material for "Uromodulin as a protein and genetic biomarker for hypertension: a detailed systematic review & meta-analysis": Table S1

**Table S1.** Complete Search Strategy using PUBMED and EMBASE. The search was performed from the date of inception until December 13, 2025.

| | |
|---|---|
| PUBMED | ("uromodulin"[Title/Abstract] OR "UMOD"[Title/Abstract] OR "Tamm-Horsfall protein"[Title/Abstract]) AND ("cardiovascular disease"[Title/Abstract] OR "CVD"[Title/Abstract] OR "coronary artery disease"[Title/Abstract] OR "CAD"[Title/Abstract] OR "ischemic heart disease"[Title/Abstract] OR "IHD"[Title/Abstract] OR "myocardial infarction"[Title/Abstract] OR "MI"[Title/Abstract] OR "heart"[Title/Abstract] OR "heart failure"[Title/Abstract] OR "atrial fibrillation"[Title/Abstract] OR "vascular disease"[Title/Abstract] OR "cardiovascular event"[Title/Abstract] OR "cardiac event"[Title/Abstract] OR "MACE"[Title/Abstract] OR cardiac[Title/Abstract] OR hypertension[Title/Abstract]) |
| EMBASE | ('uromodulin':ab,ti OR 'umod':ab,ti OR 'tamm-horsfall protein':ab,ti) AND ('cardiovascular disease':ab,ti OR 'cvd':ab,ti OR 'coronary artery disease':ab,ti OR 'cad':ab,ti OR 'ischemic heart disease':ab,ti OR 'ihd':ab,ti OR 'myocardial infarction':ab,ti OR 'heart attack':ab,ti OR 'mi':ab,ti OR 'heart':ab,ti OR 'heart failure':ab,ti OR 'atrial fibrillation':ab,ti OR 'vascular disease':ab,ti OR 'cardiovascular event':ab,ti OR 'cardiac event':ab,ti OR 'mace':ab,ti OR 'cardiac':ab,ti OR 'hypertension':ab,ti) NOT 'conference abstract'/it |
